# Genome-wide association study implicates novel loci and reveals candidate effector genes for longitudinal pediatric bone accrual through variant-to-gene mapping

**DOI:** 10.1101/2020.02.17.20024133

**Authors:** Diana L. Cousminer, Yadav Wagley, James A. Pippin, Ahmed Elhakeem, Gregory P. Way, Shana E. McCormack, Alessandra Chesi, Jonathan A. Mitchell, Joseph M. Kindler, Denis Baird, April Hartley, Laura Howe, Heidi J. Kalkwarf, Joan M. Lappe, Sumei Lu, Michelle Leonard, Matthew E. Johnson, Hakon Hakonarson, Vicente Gilsanz, John A. Shepherd, Sharon E. Oberfield, Casey S. Greene, Andrea Kelly, Deborah Lawlor, Benjamin F. Voight, Andrew D. Wells, Babette S. Zemel, Kurt Hankenson, Struan F. A. Grant

## Abstract

Bone accrual impacts lifelong skeletal health, but genetic discovery has been hampered by cross-sectional study designs and uncertainty about target effector genes. Here, we captured this dynamic phenotype by modeling longitudinal bone accrual across 11,000 bone scans followed by genome-wide association studies (GWAS). We revealed 40 loci (35 novel), half residing in topological associated domains harboring known bone genes. Variant-to-gene mapping identified contacts between GWAS loci and nearby gene promoters, and siRNA knockdown of gene expression clarified the putative effector gene at three specific loci in two osteoblast cell models. The resulting target genes highlight the cell fate decision between osteogenic and adipogenic lineages as important in normal bone accrual.

## Main text

Osteoporosis is a chronic disease characterized by low bone mineral density (BMD) and strength, which subsequently increase risk of fracture. Bone acquisition during growth is critical for achieving optimal peak bone mass in early adulthood and influences how bone density tracks throughout life^1^; individuals with higher peak bone mass ultimately have lower risk of later-life fracture^2^. Thus, understanding the factors that contribute to bone accrual has fundamental implications for optimizing skeletal health throughout life^3,4^.

Skeletal growth is a dynamic process involving bone formation driven by osteoblasts and resorption by osteoclasts. During growth, the accrual rate of areal BMD (aBMD), a measure often used to assess bone development clinically, varies by skeletal site and maturational stage^5^. aBMD is highly heritable, and while >1,000 genetic variants are associated with aBMD in adults^6–8^, much less progress has been made in identifying genetic determinants of aBMD during growth^9–11^. Although many adult-identified loci also associate with pediatric aBMD^12^, the influences of some genetic factors are principally limited to periods of high bone-turnover, such as during bone accrual in childhood^13^. However, given that pediatric genetic studies of bone accrual to date have mainly employed cross-sectional study designs, intrinsic limits are placed on the discovery of genetic variants that influence dynamic changes in bone accrual during development.

Furthermore, because the causal effector genes at many loci identified by GWAS have not yet been identified, these signals have offered limited insight without extensive follow-up. Typically, GWAS signals have been assigned to the nearest gene, but given improvements in our understanding of the spatial organization of the human genome^14^, proximity may not imply causality. As a result, variant-to-gene mapping has become an increasingly popular, evidence based approach across a range of complex traits to link association signals to target gene(s). Chromatin conformation-based techniques that detect contacts between distant regions of the genome provide one piece of evidence connecting non-coding putative regulatory sequences harboring phenotypically associated variants to a nearby gene of interest; indeed, such data are particularly powerful when there is a paucity of eQTL data for trait-relevant cell types.

Recognizing the importance of understanding the factors influencing bone accrual to maximize lifelong bone health, we leveraged ∼11,000 bone density measurements in the Bone Mineral Density in Childhood Study (BMDCS). By longitudinally modeling bone accrual in this cohort, we were subsequently well-placed to perform a series of genetic discovery analyses. Our approach implicated both putative causal variants and corresponding effector genes through the use of our variant-to-gene mapping pipeline^15^. We then further investigated specific loci to characterize their impact on osteoblast function in two relevant human cell models. Throughout the text, we describe loci based on the typical nearest gene nomenclature in order to orientate the reader, but we do not intend to imply that this gene is necessarily causal unless experimental evidence is found for that gene.

## Results

### Longitudinal modeling of aBMD and BMC

We modeled aBMD (g/cm^2^) and BMC (g/cm) from age 5 to 20 years in the BMDCS, a mixed longitudinal, multiethnic cohort of healthy children and adolescents with up to seven annual measurements. Participants were recruited to create national reference curves^16^ from five sites across the United States (**Figure 1A; Supplementary Table 1**). We modeled sex- and ancestry-specific bone accrual with ‘Super Imposition by Translation and Rotation’ (SITAR)^17^, a shape invariant model that generates a population mean curve based on all measurements. The resulting individual growth curves were then defined relative to the population mean curve by shifting in three dimensions, resulting in three random effects for each individual: *a-size:* up-down on the y-axis, representing differences in mean aBMD or BMC; *b-timing:* left-right on the x-axis, measuring differences in age when the growth rate increases; and *c-velocity*: stretched-compressed on the age scale to measure differences in the bone accrual rate (**Figure 1B**). We accessed previously derived SITAR models of BMC at the lumbar spine, total hip, femoral neck, and distal ⅓ radius^18^, and performed additional modeling for aBMD at these sites. We also modeled BMC and aBMD at the total body less head (TBLH) and skull (**Figure 1C**). Mean curves by sex and ancestry for aBMD and BMC at the six skeletal sites are shown in **Supplementary Figure 1**.

**Figure 1.**
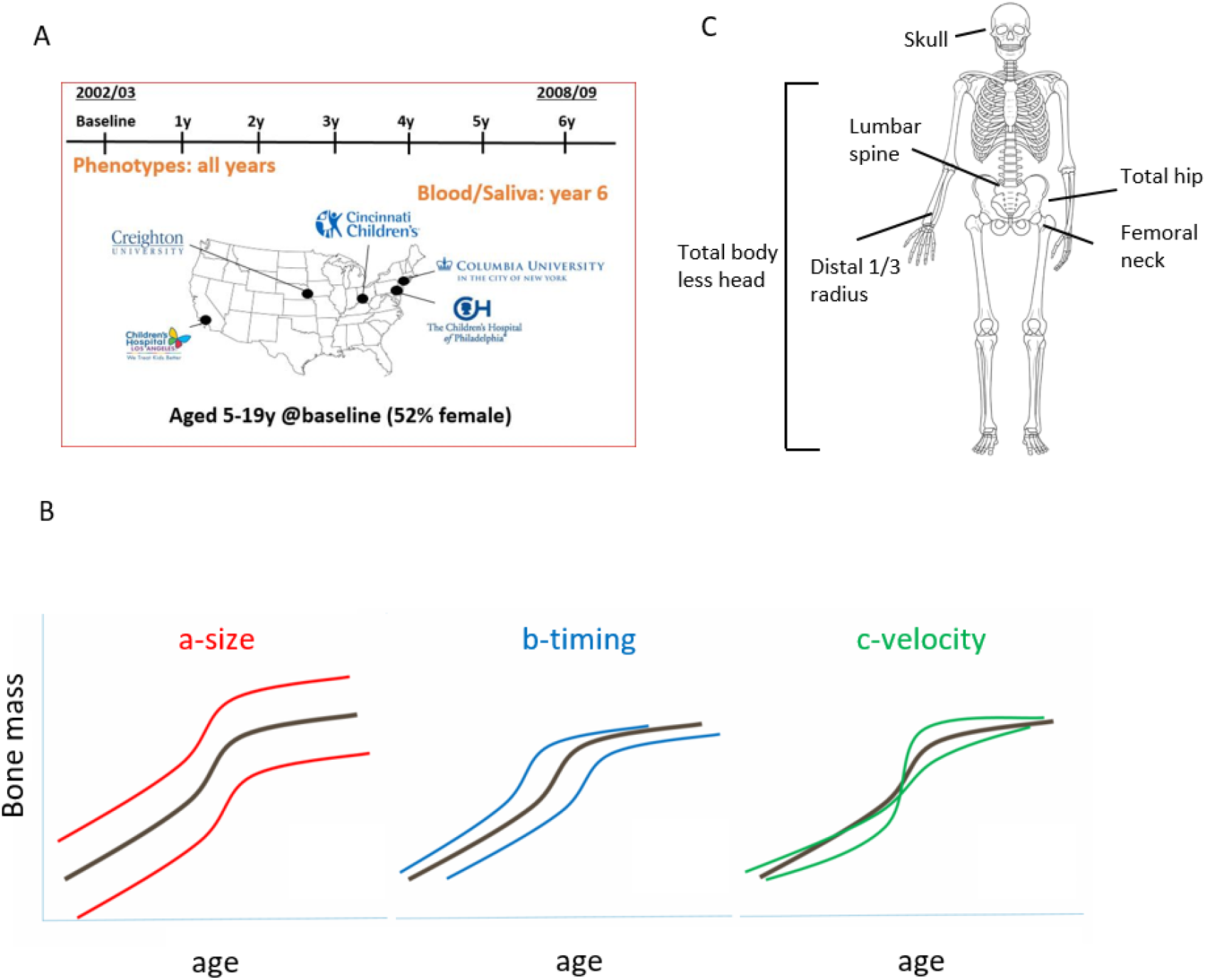
Study design and longitudinal modeling. A) Bone Mineral Density in Childhood Study (BMDCS) was a multi-ethnic longitudinal prospective study of healthy children and adolescents collected over 7 years at 5 clinical sites across the US to establish national reference curves for bone density. B) The three SITAR model parameters are a-size, representing an up-down shift on the y-axis for an individual compared to the population mean; b-timing, representing an earlier-later shift on the x-axis compared to the population mean; and c-velocity, corresponding to differences in the rate of bone accrual. C) Six skeletal sites were assessed (total body less head, 1/3 distal radius, lumbar spine, femoral neck, total hip, and skull) for bone mineral density (g/cm2) and content (g).

### Heritability of longitudinal pediatric bone density varies by skeletal site

To improve and extend heritability estimates of bone traits, we leveraged both directly genotyped and imputed variants to calculate SNP-heritability (*h*^*2*^_*SNP*_) for aBMD and BMC at the six skeletal sites (see **Supplementary Table 2** for heritability power calculations). In both cross-sectional and longitudinal analyses for the *a-size* parameter, *h*^*2*^_*SNP*_ was highest for the skull and lowest for the 1/3 distal radius (**Figure 2**; **Supplementary Tables 3 & 5**), and the results remained largely unchanged when modeling African American (AA) and non-AA participants together or separately (**Supplementary Table 4**). Therefore, for subsequent genetic analyses, we used ancestry- and sex-specific modeling results. aBMD and BMC estimates for *b-timing* varied, sometimes being substantially lower (as for the skull) or higher (as for total hip BMC) than their *a-size* counterparts. Finally, heritability estimates were overall lowest for *c-velocity*, but had a larger range, from *h*^*2*^_*SNP*_ (SE) = 0.092 (0.089), *P* = 0.15 for distal radius aBMD to *h*^*2*^_*SNP*_ (SE) = 0.69 (0.088), *P* = 2.51×10^−13^ for skull BMC. These results show that *h*^*2*^_*SNP*_ was robust across ancestry groups, encouraging us to consider AA and non-AA participants jointly for genetic discovery efforts, and that each of the three SITAR parameters displayed a significant genetic component across skeletal sites.

**Figure 2.**
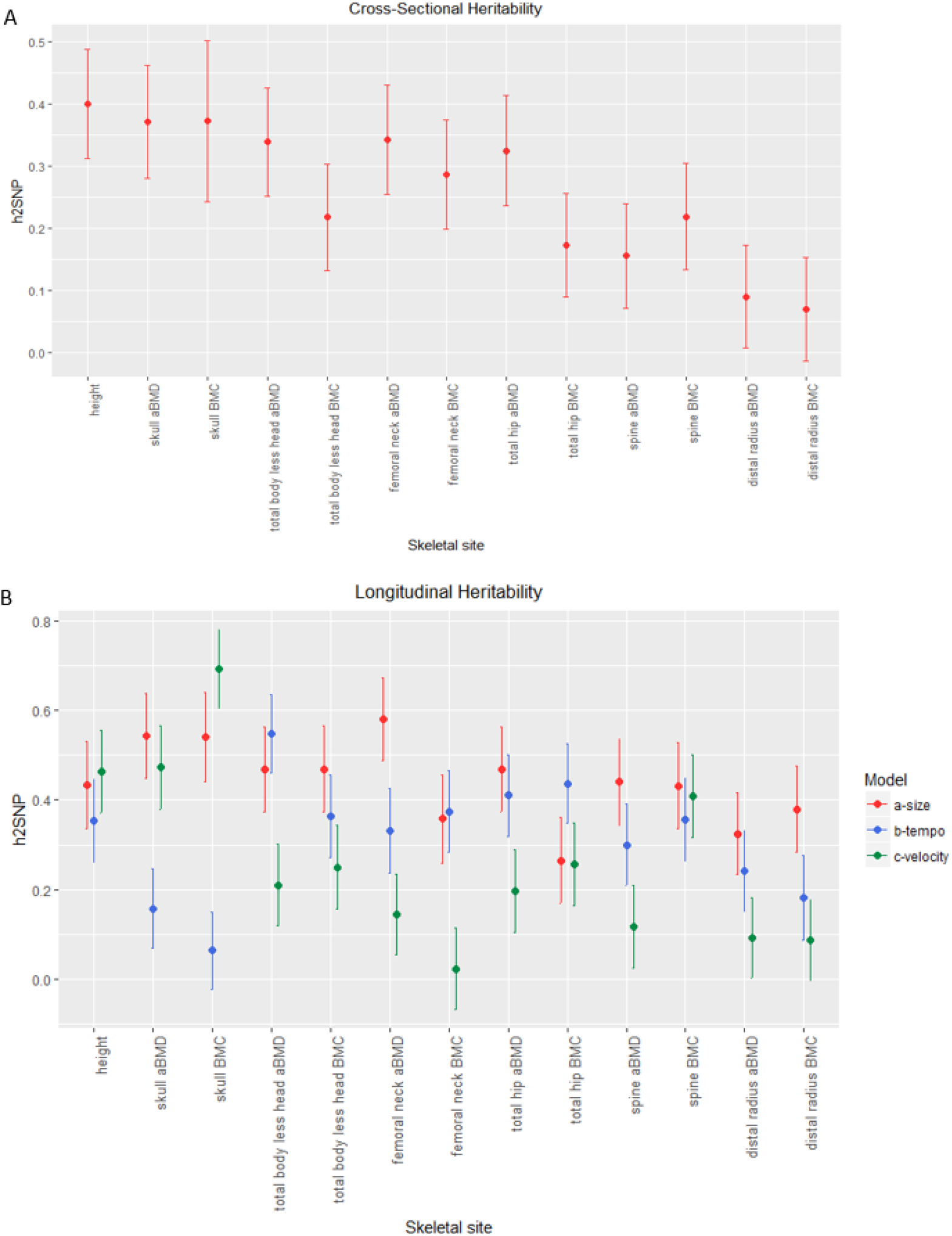
SNP-based heritability estimates. A) Estimates of heritability for cross-sectional baseline data. B) Heritability estimates derived from longitudinal growth modeling with SITAR. Heritability estimates, standard errors, and P-values are given in Supplementary tables 3 and 5.

### GWAS reveals novel loci associated with pediatric bone accrual

Next, to identify loci associated with bone accrual, we performed 36 parallel GWAS on the three SITAR parameters (*a-size, b-timing, c-velocity*) for aBMD and BMC at the six skeletal sites (**Supplementary Figure 2)**. Twenty-seven association signals achieved the traditional genome-wide significance threshold of *P* < 5×10^−8^, with many associated with more than one skeletal site or parameter (designated as signals 1-27, ordered by chromosome and position; **Table 1**). Acknowledging the large number of statistical tests performed, we used several strategies to prioritize loci for further analyses. First, given the high correlation between aBMD and BMC and among different skeletal sites (**Supplementary Figure 3; Supplementary Tables 6 and 7**), we used PhenoSpD^19,20^ to calculate the number of independent tests. This revealed an equivalent of 16 independent tests, resulting in a corrected significance threshold of *P* < 3.1×10^−9^, which yielded one locus (signal 26, rs201392388, nearest gene *FGF16*) surpassing the corrected genome-wide significance threshold accounting for multiple testing. Second, ten loci achieved a suggestive significance level (*P* = 5×10^−8^ - 1×10^−6^) and were supported by more than one of our phenotypes (designated signals S1-S10). Finally, we set aside three loci that reached suggestive significance in one phenotype but also gained support (*P* < 10^−4^) from a recent GWAS of adult heel eBMD in the UK Biobank^6^ (designated signals S11-S13). This brought the total number of prioritized loci for follow-up assessment to 40. Overall, most loci yielded similar effect sizes in males and females and in both ancestry groups (**Supplementary Table 8**). Only one of these loci was previously associated with pediatric aBMD (signal 16, rs17140801^9^, nearest gene *RBFOX1*). In addition to the suggestive signals S11-S13, three other signals were associated with adult heel eBMD (**Supplementary Table 10**), and one signal was previously associated with adult lumbar spine aBMD^7^. In total, 35/40 (87.5%) of our signals were novel.

**Table 1.** Genome-wide significant loci (signals 1-27) and suggestive loci (signals S1-S13) supported by multiple phenotypes or skeletal sites.

### Follow-up in ALSPAC

We attempted replication of loci in the ALSPAC cohort (n=6,382), but were limited by the skeletal sites available. TBLH and skull BMC were modeled with SITAR, after which the *a-size, b-timing*, and *c-velocity* random effects were subjected to GWAS using a linear mixed model. Given the differences between the BMDCS and ALSPAC data (including different DXA machines used, populations (mixed ancestry US vs. British white), ages (beginning at age 5yrs in BMDCS and 10yrs in ALSPAC), cohort-specific covariates applied, and genotyping arrays employed), we opted to take a broad approach and extracted all SNPs in LD with our 40 lead signals (r^2^ > 0.8 in Europeans). This analysis provided support for five of our loci (**Supplementary Table 9**), one of which also showed suggestive support in the heel eBMD lookup (signal S11, rs2564086, nearest gene *SOX11*).

### Association with later-life fracture in adults

Recently, we found that the postmenopausal bone loss and fracture-associated Sp1 variant within the *COLIA1* gene^1,2^ was implicated in delayed bone gain across puberty in girls^13^. Given that both bone gain and loss are periods of relatively high bone turnover, we assessed the converse possibility: that bone accrual-associated variants might also influence later-life fracture risk. We queried our signals in a published UK Biobank GWAS of adult fracture^6^ and found that three loci showed at least nominal association (*P* < 0.05) with fracture risk (**Supplementary Table 10**), including one of our suggestive signals that was associated with both heel eBMD and fracture (signal S13, rs11195210, nearest gene *SMC3*; heel eBMD beta =-0.02, *P* = 2.3×10^−8^; fracture OR = 1.04, *P* = 0.0024). For eight loci, we found additional support for association with fracture phenotypes in the UK Biobank using the PheWeb browser (**Supplementary Table 11**), including signal 22 (rs8130725, nearest gene *NRIP1*), which was associated with self-reported fracture of the radius (*P* = 8.9×10^−5^; surpassing a corrected significance threshold of 1.25×10^−3^). Thus, we identified loci putatively active in bone metabolism both early and later in life.

### Functionally relevant candidate genes and pathways at associated loci

We next sought to identify credible candidate effector genes near the 40 prioritized loci. Given that the nearest gene to a GWAS signal is often not the causal gene, we considered all genes within the signals’ surrounding topological associated domains (TADs), regions of the genome previously defined as most likely to set the bounds where the causal effector gene resides^21^. This resulted in 319 protein-coding genes (all TAD genes are listed in **Supplementary Table 12**). We then assessed the extent of evidence supporting that this set of genes is involved in skeletal development. At 20 loci (with two harboring two distinct signals each), we observed genes known to be involved in bone biology (**Table 2; Supplementary Table 13**). Many of these genes are well-established as key players in osteogenesis or skeletal development, such as *FOSL2*, which controls osteoclast size and survival^22^; *WWTR1* (*TAZ*), encoding a key member of the Hippo pathway that interacts with RUNX2 to induce osteogenesis^23^; *SLC9A3R1* (*NHERF*), a member of the Wnt signaling pathway associated with hypophosphatemic nephrolithiasis/osteoporosis-2 (OMIM 612287) as well as low bone mineral density^24^; and *TGFB1*, mutations in which lead to Camurati-Englelmann disease (OMIM 131300) and bone density alterations^25^. We also observed important skeletal biology-related genes at suggestive loci, with these genes including *TWIST2* (Ablepharon-macrostomia syndrome and Barber-Say syndrome [OMIM 200110 & 209885, respectively], which show premature osteoblast differentiation and growth retardation^26–28^); *HDAC4* (‘osteoblast differentiation and development’)^29,30^; *PRKD1* (‘positive regulation of osteoclast development’)^31^; *HMG20B* (‘osteoblast differentiation and development’); and *SOX11* (Coffin-Siris syndrome 9 [OMIM 615866] in which there is abnormal skeletal morphology)^32^. Although these are known genes, we note that genetic associations have not been previously implicated nearby these genes in GWAS of aBMD and BMC (with the exception of the *TGFB1* locus^6^). This analysis revealed plausible candidate effector genes at half of the association signals, although direct evidence linking our signals to these genes remains to be established.

**Table 2.** Candidate genes with evidence for functional involvement in skeletal biology.

| Signal | Gene<sup>a</sup> | Gene Ontology or KEGG term | Mendelian disease (OMIM ID)<sup>b</sup> | Human phenotype | Mouse phenotype<sup>c</sup> | Selected references |
| --- | --- | --- | --- | --- | --- | --- |
| 2 | *FOSL2* | Osteoclast differentiation | NA | NA | abnormal osteoblast and osteoclast morphology and physiology; abnormal bone morphology; decreased bone mineralization; etc. | Bozec (*Nature* 2008) |
| 3 | *FOXP1* | Osteoclast differentiation & development | NA | NA | abnormal osteoclastogenesis and bone resorption | Zhao (*Dev Biol* 2015) |
| 5 | *WWTR1*<br>(*TAZ*) | Osteoclast differentiation, mesenchymal cell differentiation | NA | NA | abnormal skeleton morphology & bone ossification; decreased body size | Hong (*Science* 2005) |
| 8 | *IL17A* [P] | Positive regulation of osteoclast development | NA | NA | NA | Kotake (*Clin Invest* 1999) |
| 9 | *TBXAS1* | NA | Ghosal heamtodiaphyseal dysplasia (231095) | increased bone density | NA | Genevieve (*Nat Genet* 2008) |
| 15 | *BLNK* | Osteoclast differentiation | NA | NA | NA | Shinohara (*Cell* 2008) |
| 18 | *SLC9A3R1*<br>(*NHERF*) | Wnt signaling pathway | hypophosphatemic nephrolithiasis/osteoporosis-2 (612287) | low bone mineral density | decreased bone mineral content and density | Karim (*NEJM* 2008) |
| 19 | *GRB2* [P] | Osteoclast differentiation | NA | NA | abnormal mandible morphology | Levy-Apter (*J Biol Chem* 2014) |
| 21 | *TGFB1* | Osteoclast differentiation | Camurati-Engelmann disease (131300) | cortical thickening of the diaphyses of the long bones; some patients also show involvement of the skull; increased bone mineral density; osteopenia | abnormal bone ossification; decreased osteoblast cell number; etc. | Kinoshita (*Nat Genet* 2000); Gao (*Proc Natl Acad Sci* 2004) |
| 22-23 | *NRIP1* | NA | NA | Differentially expressed in low and high BMD samples; may interact with ESR1 | NA | Morón (*Bone* 2006); Li (*Bone Joint Res* 2016) |
| 24 | *CITED1* | Negative regulation of osteoblast differentiation | NA | NA | NA | Yang (*Endocrinology* 2008) |
| 25-26 | *ATP7A* | NA | Menkes disease (309400), Occipital horn syndrome (304150) | defective collagen cross-linking resulting in osteoporosis and pathological fracture | abnormal skeleton morphology; osteoarthritis | Kim (*Stem Cell Res Ther* 2015) |
| S2 | *HDAC4* | Osteoblast differentiation & development | NA | NA | abnormal bone ossification; abnormal osteoblast cell number; skeletal phenotype; etc. | Vega (*Cell* 2004) |
|  | *TWIST2* | Negative regulation of osteoblast differentiation | Ablepharon-macrostomia syndrome (200110); Barber-Say syndrome (209885) | premature osteoblast differentiation, growth retardation | abnormal osteoblast differentiation | Bialek (*Dev Cell* 2004) |
| S3 | *TET2* [P] | NA | Myelodysplastic syndrome, somatic (614286) | NA | Osteopetrosis, reduced osteoclast number | Chu (*Genomics, Proteomics & Bioinf.* 2018); Yang (*Nat Comm* 2018) |
| S6 | *TEAD4* | Skeletal system development | NA | NA | NA | Matsumoto (*J Clin Invest* 2016); |
|  | *TULP3* [P] | Bone development | NA | NA | abnormal vertebrae morphology & development | Patterson (*HMG* 2009) |
| S7 | *PRKD1* | Positive regulation of osteoclast development | NA | NA | reduced bone mineral density; decreased trabecular thickness | Bollag (*Mol Cell Endocrinol* 2018) |
| S9 | *GNA11* | Skeletal system development | Hypocalcemia (615361); hypocalciuric hypercalcemia (145981) | altered calcium levels; short stature | decreased bone mineral content and density | Nesbit (*NEJM* 2013) |
| S9 | *AES* (*TLE5*) | Skeletal system development | NA | NA | NA | Zhao (*BBRC* 2017) |
| S11 | SOX11 | Positive regulation of osteoblast differentiation | Coffin-Siris syndrome 9 (615866) | abnormal skeletal morphology | abnormal bone mineralization & ossification; abnormal lumbar vertebrae morphology | Tsurusaki (*Nat Commun* 2014) |
| S12 | SLIT3 | NA | NA | higher circulating levels associated with higher bone mass in postmenopausal women | low bone mass | Kim (*J Clin Invest* 2018) |
| S13 | SMC3 | NA | Cornelia de Lange syndrome 3 (610759) | growth retardation | decreased bone mineral content | Andrade (*Horm Res Paediatr* 2017) |

<sup>a</sup>[P] Genes implicated by promoter-interaction map
<sup>b</sup>Mendelian disease genes from the Online Mendelian Inheritance in Man (OMIM) database (https://www.ncbi.nlm.nih.gov/omim)
<sup>c</sup>Mouse phenotypes from the Mouse Genome Informatics database (http://www.informatics.jax.org/)

Next, we performed pathway analysis for all transcripts in the TADs corresponding to the 40 prioritized signals,^33^ which revealed several pathways of interest, including ‘long-chain fatty acid metabolic process’, ‘negative regulation of toll-like receptor signaling pathway’, ‘Calcium signaling pathway’, ‘FoxO signaling pathway’, and ‘Hippo signaling pathway’ (**Supplementary tables 14 & 15**).

### Variant-to-gene mapping identifies high-confidence SNP-to-gene promoter contacts

We then performed variant-to-gene mapping to physically connect our signals with their putative target effector genes (overview of analytical pipeline provided in **Figure 3A**). In order to implicate effector genes in an appropriate tissue context, we leveraged data from human mesenchymal stem cell (hMSC)-derived osteoblasts^15^. We first extracted all proxy SNPs in LD with our lead SNPs (liberal r^2^ ≥ 0.5) that resided in accessible chromatin^15^. Next, we queried accessible SNP-to-gene interactions in high-resolution promoter-focused Capture C data from the same cell line. Six loci (15% of the 40 loci identified) exhibited *cis* interactions with gene promoters (**Supplementary Table 16**), with a total of 22 genes targeted by these interactions. These target genes included several prioritized by our functional candidate search, such as *GRB2* (signal 19), involved in osteoclast differentiation^34^ and *TULP3* (signal S6), associated with abnormal vertebrae morphology and development^35^.

**Figure 3.**
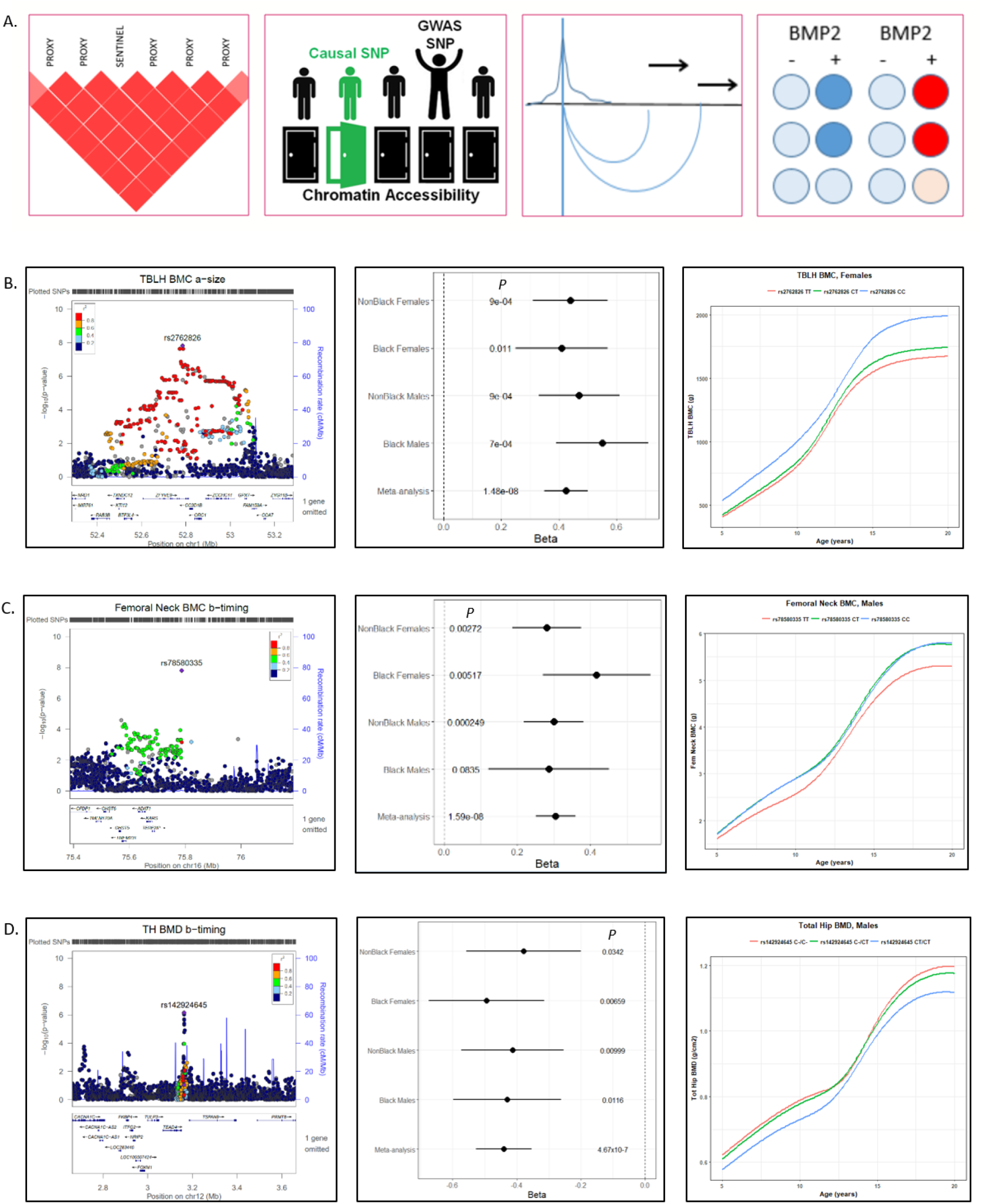
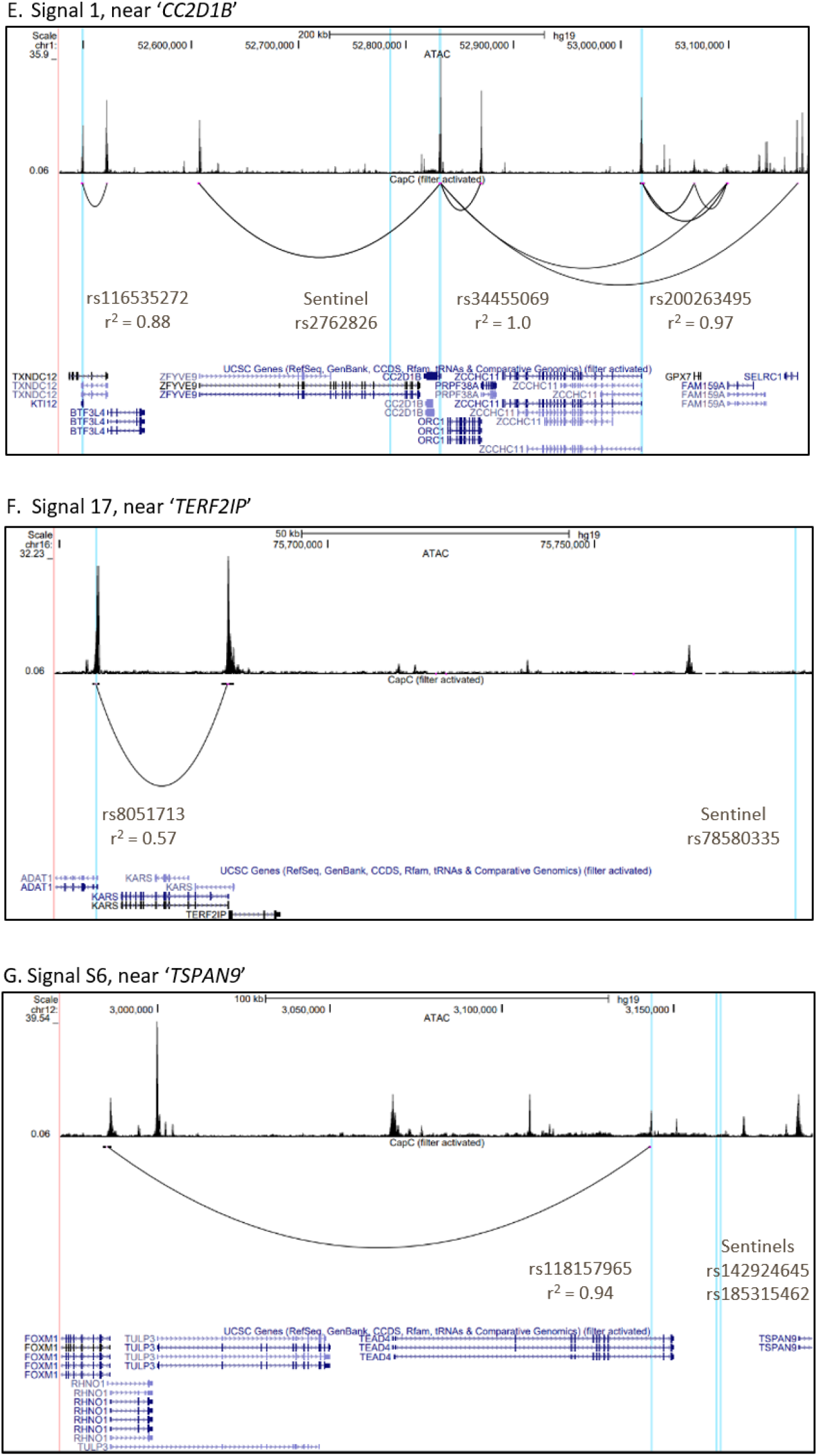
Genome-wide association and variant-to-gene mapping highlight three loci associated with pediatric bone accrual. A) Overview of variant-to-gene mapping pipeline. B-D) Locuszoom plots, association results in each subsample (Black and Non-Black males and females), and representative mean SITAR curves by genotype for (B) signal 1, near ‘*CC2D1B*’, (C) signal 17, near ‘*TERF2IP*’, and (D) signal S6, near ‘*TSPAN9*.’ E-G) UCSC Genome Browser images for these three loci, showing the sentinel and proxy SNPs in vertical blue lines. The top panel of each browser image shows open chromatin peaks assessed by ATAC-seq, with *cis* interaction loops shown below.

Our promoter-focused Capture C data also pointed to the nearest gene at signal S3, *TET2*, a known promoter of osteogenesis^36^ (**Supplementary Figure 4A**). Interestingly, we observed two association signals at this locus, which despite being in moderate LD (r^2^ = 0.43) showed opposite directional effects (**Table 1**). These two signals were both genome-wide significant in the published heel eBMD GWAS^6^ with opposite effect directions (**Supplementary Table 10**). One of our signals, rs56883672-C (FN BMC c-velocity, beta (SE) = 0.21 (0.038), *P* = 7.1×10^−8^) was in LD with an open proxy SNP, rs2301718, which showed a *cis* interaction with *TET2* and falls in a binding site for RBPj, a primary nuclear mediator of Notch and an osteogenic driver^37^ (**Supplementary Figure 4B**). Thus, rs2301718 is a putative causal variant falling in a potential novel distal enhancer for *TET2*. Additionally, a conserved binding site for FOXO3 (determined by the Transfac Matrix Database (v7.0) in the UCSC Genome Browser), a transcription factor regulated by RBPj^38^ and a member of the FOXO family which play critical roles in skeletal homeostasis^39^, lies immediately upstream (**Supplementary Figure 4C**). Thus, this may be a regulatory region for osteogenesis with dynamic effects at different skeletal sites.

Although GTEx data was not generated for bone, we searched for pan-tissue eQTLs that would provide evidence linking our loci to effector genes. We identified SNPs in high LD (r^2^ > 0.8) with the 40 sentinel SNPs, queried significant eQTLs in all available GTEx tissues^40^ and observed eQTLs for 28 genes (**Supplementary Table 17**). None of the genes highlighted by our functional search overlapped with eQTL genes, suggesting tissue-specific or temporal-specific regulation that is not reflected in this broad pan-tissue context.

### Functional assays in bone cell lines implicates one novel gene each at three loci

About half of our prioritized loci lacked clear functional candidate effector genes. Two of these loci (signals 1 and 17, near *CC2D1B* and *TERF2IP*, respectively) proved more challenging to resolve given they both had multiple gene contacts in our hMSC-derived osteoblast atlas (**Figure 3 B**,**C**,**E**,**F**) as well as pan-tissue eQTL support. To identify genes involved in bone mineralization, we followed up putative candidate effector genes identified by eQTLs and/or variant-to-gene mapping at these two loci. Another locus had support for more than one plausible candidate effector gene, so we also aimed to clarify this observation (signal S6; **Figure 3 D**,**G**). We performed siRNA knockdown of four genes at each locus (for a total of 12 genes) in primary hMSCs and assessed osteoblast differentiation. qPCR analysis revealed that each siRNA resulted in specific, significant knockdown of its corresponding target under unstimulated conditions (**Supplementary Figure 5**).

To identify which contacted genes have roles in osteoblast function, we examined osteoblast activity with histochemical alkaline phosphatase (ALP) and mineralization with Alizarin red S staining. We found that disruption of just one gene per locus among each group of four candidates showed a significant reduction in terminal osteoblast differentiation. While targeting *GPX7, PRPF38A, ORC1*, or *ZCCHC11* at signal 1 produced somewhat variable ALP staining across donor lines, the staining levels (**Figure 4A-B**) and corresponding *ALPL* gene expression levels (**Figure 4C**) were not significantly different from non-targeted cells. On the other hand, there was a marked reduction in Alizarin red S staining after *PRPF38A* knockdown (**Figure 4A and D**).

**Figure 4.**
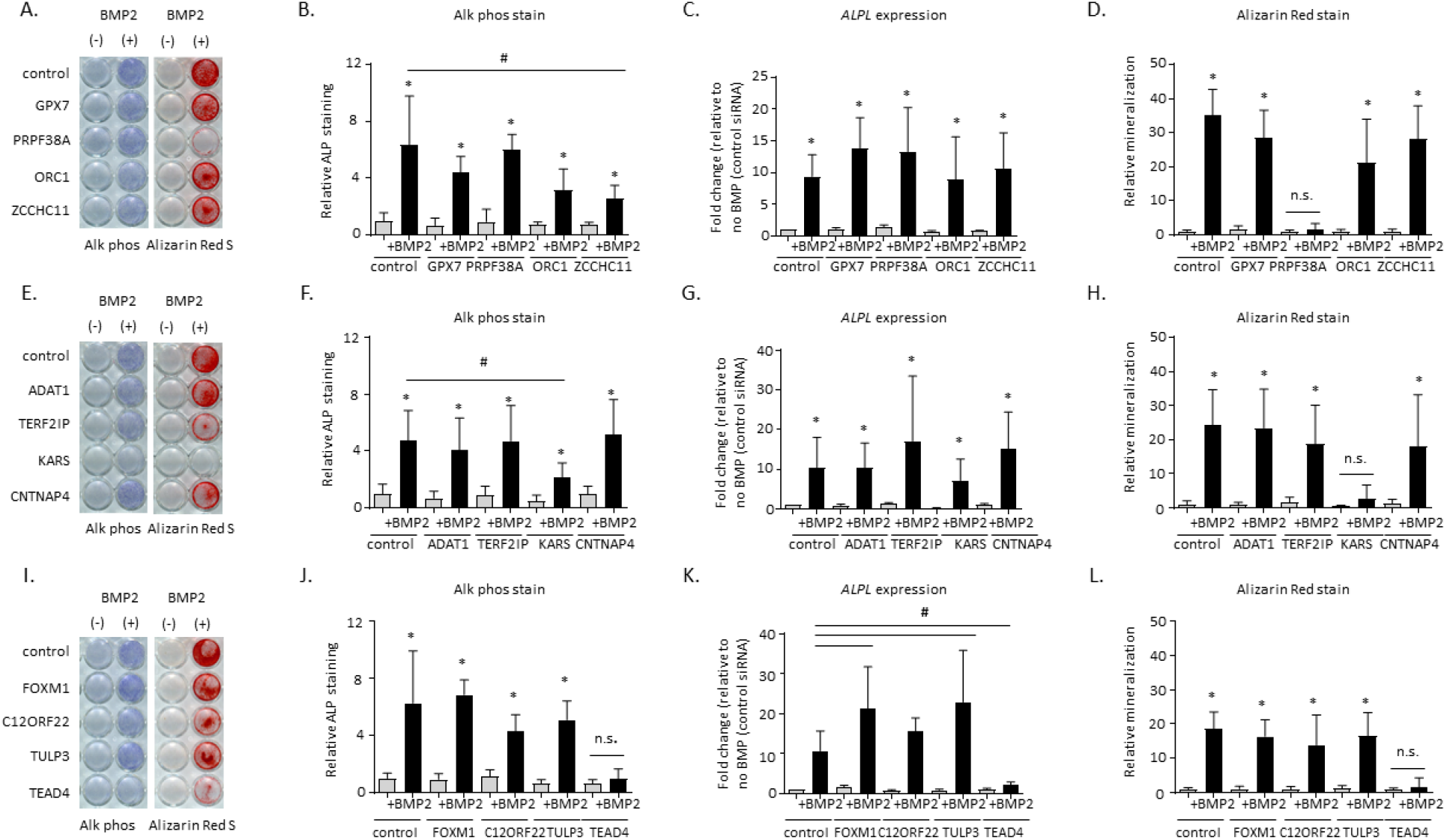
Functional assays in human mesenchymal stem cell-induced osteoblasts following siRNA knockdown of four genes each at three loci implicated by GWAS and variant-to-gene mapping. A, E, I) Representative alkaline phosphatase (blue) and Alizarin Red S (red) stains for osteoblastic activity and calcium deposition, respectively, for the three tested loci. Experiments were performed twice in three unique donor lines. B, F, J) Quantification of Alkaline phosphatase staining using quantitative image analysis was repeated twice with three different independent hMSC donor cell lines. C, G, K) *ALPL* gene expression. D, H, L) Quantification of Alizarin Red S staining. * *p*<0.05 comparing No treatment to BMP treatment for each siRNA, #*p*<0.05 comparing control siRNA to siRNA for gene of interest, n.s = not significant. Error bars represent standard deviation.

At the signal 17 locus (targeting *ADAT1, TERF2IP, KARS*, and *CNTNAP4*), downregulation of *KARS* produced a significant reduction in ALP staining and extracellular calcium deposition, but we did not observe a significant reduction in *ALPL* gene expression itself (**Figure 4E-H**). To further understand the discrepancy between ALP staining and gene expression patterns, we individually examined the ALP expression and staining pattern for each donor line. A consistent reduction in *ALPL* gene expression and ALP staining was clearly evident in two out of the three donor lines, but despite a reduction of ALP staining in the third line, *ALPL* gene expression was increased (data not shown). Despite the donor variability seen in our experiments, male mice with a heterozygous *KARS* knockout have a significant increase in BMD excluding skull (male *P* = 6.61×10^−5^; significance threshold 1×10^−4^; <https://www.mousephenotype.org/), >providing orthogonal evidence for the importance of this gene in osteogenesis.

At signal S6 (targeting *FOXM1, C12ORF22, TULP3*, and *TEAD4*), although there are several plausible functional candidate genes (**Table 2**), targeting *TEAD4* significantly reduced both ALP staining and *ALPL* gene expression as well as extracellular calcium deposition (**Figure 4I-L**). In contrast, gene knockdowns for the other candidates had no impact on these readouts. These results were subsequently replicated in siRNA knockdown experiments in an immortalized human fetal osteoblast (hFOB) cell line (**Supplementary Figure 6**).

Activation of canonical BMP signaling leads to the phosphorylation of SMAD proteins and upregulation of the *ID1* family of genes. Thus, we assessed BMP2 signaling by measuring *ID1* gene expression and assessed expression of pro-osteoblastic transcription factors *RUNX2* and *SP7*. After *PRPF38A* and *KARS* knockdown, BMP2 signaling was intact and the expression of *RUNX2* and *SP7* were preserved after knockdown of *PRPF38A* and *KARS* (**Supplementary Figure 7A-F**). For cells lacking *TEAD4, ID1* expression was unchanged, although *RUNX2* and *SP7* expression levels were lower than observed in controls (**Supplementary Figure 7G-I**). These results suggest that these three genes impact osteogenesis via distinct molecular mechanisms.

### PRPF38A knockdown induced morphological changes

At signal 1, *PRPF38A* silencing resulted in a morphological change and reduction of extracellular calcium deposition that was evident in both hMSC-derived osteoblasts (**Figure 5A**) and hFOBs (**Figure 5B**). Thus, we examined whether *PRPF38A* silencing affected the expression of chondrocyte specific genes *ACAN, COMP*, and *SOX9* or the expression levels of later osteoblastic genes *IBSP* and *OMD* in hMSCs. Despite some variability in our observations, our results largely showed that neither chondrocyte lineage genes nor later osteoblast specific genes were greatly altered in *PRPF38A* silenced cells (**Figure 5C-E; Supplementary Figure 8A-C**). In contrast, our results for *PRPF38A* silencing in the context of the expression of adipogenic-specific genes was more striking. *PRPF38A* silencing was sufficient to increase expression of *PPARG*, a critical transcription factor for adipocyte differentiation, and its expression increased further upon stimulation with BMP2 in hMSC-derived osteoblasts and recapitulated in hFOBs (**Figure 5F**,**G**). Likewise, *FABP4* significantly increased in *PRPF38A* silenced cells (**Figure 5H**). However, *C/EBPA* expression did not change dramatically (**Supplementary Figure 8D**). We did not observe morphological differences in the *KARS* or *TEAD4* silenced donor lines (data not shown).

**Figure 5.**
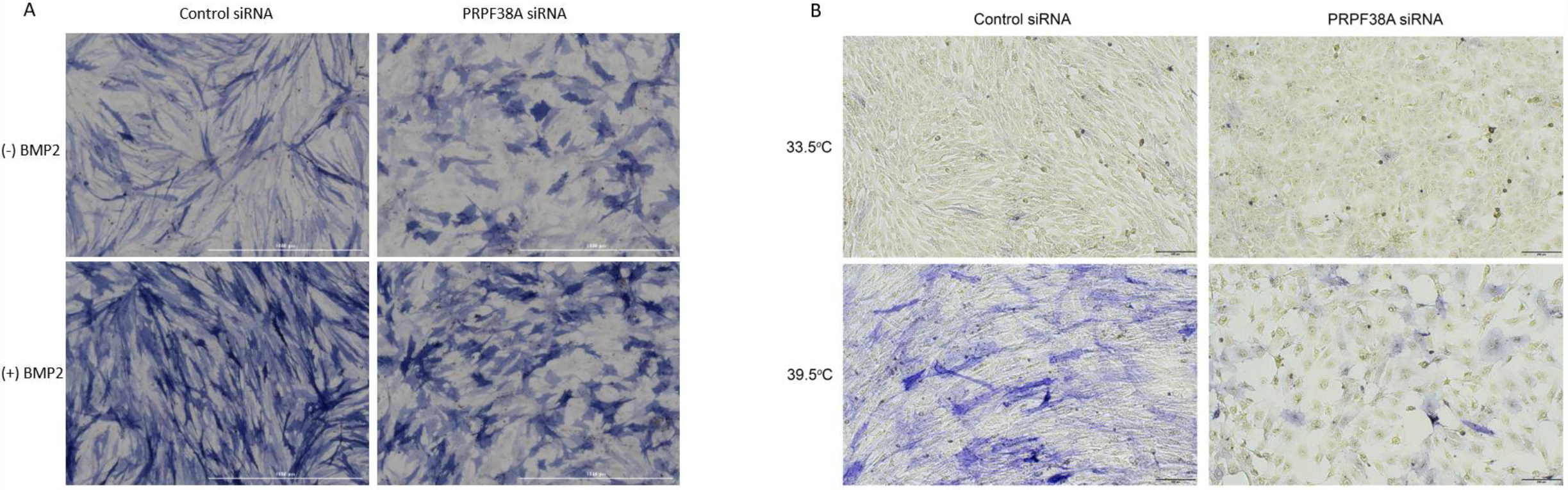
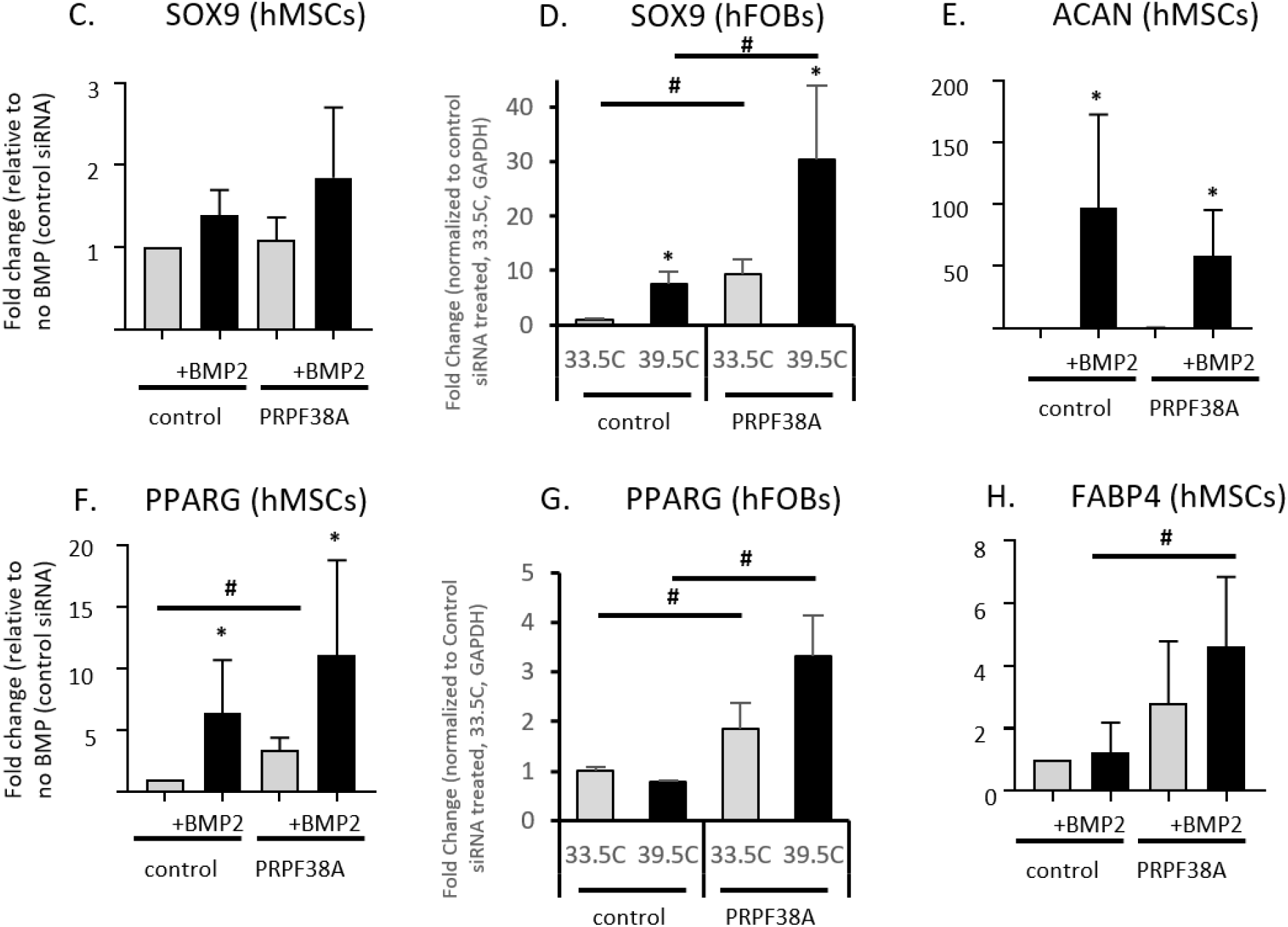
*PRPF38A* knockdown induced a morphological change in two osteoblast cell models. A) Cell morphology in hMSCs before (top) and after BMP2 treatment (bottom) with control siRNA (left) and *PRPF38A* knockdown (right). Representative color bright-field images of a typical alkaline phosphatase stained plate from *PRPF38A* silenced cells is shown. Similar morphological changes were observed for all three donor lines used in the study. Scale bar, 1000 µm. B) *PRPF38A* knockdown-induced morphological changes were recapitulated in human fetal osteoblasts under permissive growth (33.5°C; top) and differentiation (39.5°C; bottom) conditions. Scale bar, 200 µm. C-E) Quantitative gene expression of chondrocytic genes *SOX9* and *ACAN* and F-H) adipocyte-specific genes *PPARG* and *FABP4*. For hMSCs, data is from two technical replicates from three unique donor lines were averaged. For hFOBs, three technical replicates were averaged. Levels of *ACAN* and *FABP4* were undetectable in hFOBs, even in reactions with up to 600ng of cDNA (twice the amount used for qPCR of other targets). * *p*<0.05 comparing non-treated to treated cells (BMP2 or 39.5°C for hMSCs and hFOBs, respectively) for each siRNA, #*p*<0.05 comparing control siRNA to siRNA for gene of interest. Error bars represent standard deviation.

### CRISPR-Cas9 deletion of putative enhancer element for PRPF38A expression

Given the evidence for *PRPF38A* as a novel factor influencing osteogenesis in the two bone cell models, we next performed CRISPR-Cas9 deletions in hFOBs to confirm the accessible proxy SNP (rs34455069) resides within an enhancer impacting the expression of *PRPF38A*. Deletion of 123-533bp encompassing rs34455069 resulted in a 38% decrease in ALP staining (*P*=0.005, compared to empty vector control; **Figure 6A-B**) and a 45% decrease in *PRPF38A* expression (*P*=0.0009, compared to empty vector control; **Figure 6C**) as measured by qPCR. These results were obtained in a mixed cell population of wild-type and CRISPR cells (7-37% wild type) after differentiation at 39.5°C for 3 or 7 days, respectively. No morphological changes were observed in the proxy-SNP edited cells. We also deleted a 733-1823bp region around the sentinel GWAS SNP (rs2762826); as expected, perturbing the immediate region harboring the sentinel GWAS SNP had no effect on ALP staining (**Figure 6A-B**) or cell morphology. Intriguingly, rs34455069 is predicted as “likely to affect binding” (RegulomeDB; regulomedb.org) of two transcription factors with known regulatory effects in osteogenesis, KROX^41^ and SP1:SP3^42,43^ (position weight matrices for these binding sites highlighting this SNP are given in **Supplementary Figure 9**). Further work is warranted to confirm that this genetic variant, a single base-pair indel, results in differential binding of these transcription factors and altered gene expression of *PRPF38A*.

**Figure 6.**
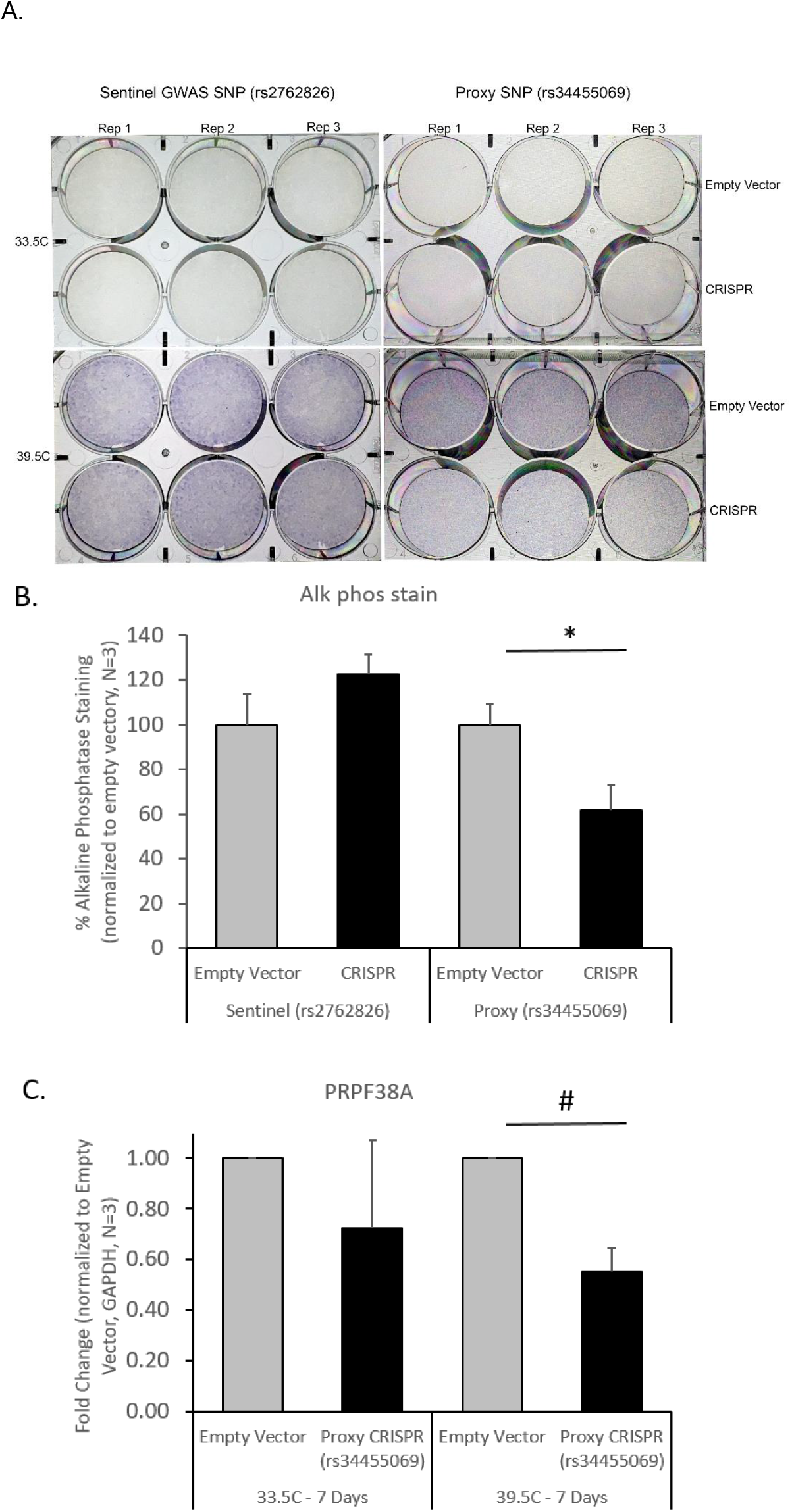
CRISPR-Cas9 deletion of sentinel and proxy SNPs at *PRPF38A* locus in hFOB cells showed that only modulation of the proxy SNP impacts alkaline phosphatase level and expression of the gene. A) Alkaline phosphatase staining was performed in triplicate after excision of the sentinel GWAS SNP (rs2762826; left) and the proxy SNP (rs34455069; right). B) Quantification of Alkaline phosphatase staining using quantitative image analysis showed that staining was reduced after excision of the region surrounding the proxy SNP, but not the sentinel SNP. C) Gene expression of *PRPF38A* was reduced after excision of the proxy SNP. * *p*<0.01, #*p*<0.001, comparing empty vector to CRISPR cells (averaged across three technical replicates). Error bars represent standard deviation.

## Discussion

To complement cross-sectional genetic studies of bone trait measurements, and to target the most dynamic changes in the trajectory of changes in bone density, i.e. during childhood, we performed longitudinal modeling to harness variation in bone mineral accrual trajectories. To assess the genetic contribution to these traits, we performed heritability analyses and GWAS, identifying 40 loci, which triples the number of identified pediatric aBMD and BMC-associated loci. Our efforts to map target genes via Capture-C and eQTL data provided implicated leads for several putative effector genes at associated loci, and we functionally characterized selected genes in two osteoblast cell models, revealing three key candidate effector genes for further study. Thus, our longitudinal approach not only revealed novel associations with pediatric bone accrual, but also the most likely functional target genes.

Focusing first on bioinformatic characterization of the 40 prioritized loci, at half of these we identified known bone-related genes residing within the corresponding TADs. Among these, WWTR1/TAZ, HDAC4, TWIST2, and PRKD1 are known to interact with RUNX2, an essential osteoblastic differentiation factor. Several genes harbor Mendelian mutations resulting in abnormal bone density or skeletal phenotypes^22–26,34,35,44–59^, and many also show abnormal mouse skeletal phenotypes (**Table 2**). Although further work is needed to concretely link many of these GWAS-implicated variants to their corresponding target effector genes, our promoter Capture-C approach did corroborate some of these genes as putative functional effector genes acting in bone accrual.

In previous work, we noted a genetic variant principally active during periods of high turnover at *COLIA1*, with implications for both delayed bone accrual in girls during puberty^13^ and post-menopausal bone loss and fracture^60,61^. Three of our signals showed evidence of association in a published GWAS of adult fracture^6^ and other fracture traits on the PheWEB browser. At signals S13 and 22, we also noted candidate genes with literature support: the nearest gene at signal S13 is *SMC3*, underlying Cornelia de Lange syndrome 3 (OMIM 610759)^62^ and decreased BMC in mice, and the nearest gene to signal 22 is *NRIP1*, which is differentially expressed in patients with low vs high BMD^49^. Thus, the *COLIA1* locus is unlikely to be the only factor influencing both bone gain and loss, and further investigation of the gene targets at these loci may provide leads to maximizing lifelong bone health.

Including genes at signals associated with various skeletal sites and parameters, pathway analysis pointed toward pathways known to be involved in bone metabolism. Several pathways were involved in long-chain fatty acid metabolism, largely driven by a cluster of cytochrome P450 (CYP) genes at a single locus (signal 21), which also harbors *TGFB1*. Although *TGFB1* is a plausible candidate gene, CYP genes metabolize eicosanoids (long-chain fatty acids) including arachidonic acid and affect metabolite levels^63^, and genetic variation in related genes (*CYP-17* and *19*) was associated with serum sex steroid concentrations and BMD, osteoporosis, and fracture in post-menopausal women^64^. Studies have shown that osteoblasts take up and metabolize fatty acids for matrix production and mineralization^65^, and long-chain fatty acids were associated with peak aBMD and bone accrual in late-adolescent males^66^.

Three loci harboring five genes (*PTPRS, CD300A, CACTIN, CD300LF, TICAM1*), were annotated with the GO term ‘negative regulation of toll-like receptor signaling pathway’. The toll-like receptor pathway has multifaceted roles in osteoblast function, including mediating bone inflammatory responses and regulating cell viability, proliferation, and osteoblast-mediated osteoclastogenesis^67^. Finally, the ‘Calcium signaling pathway’, the ‘FoxO signaling pathway’^68,69^ and the ‘Hippo signaling pathway’^70^ are fundamental in normal skeletal development.

Notably, several candidate genes are involved in TH17 cell differentiation (*IL17F, IL17A, RXRA*, and *TGFB1*). *IL17A* is a T cell derived growth factor for MSCs^71,72^, and we observed an open proxy variant contact with the *IL17A* promoter in TH17 cells but not in osteoblasts (**Supplementary Table 18**), supporting *IL17A* as the effector gene at this locus. Expression of *IL17A* inhibits the osteogenic differentiation of MSCs^73,74^, and T cell derived IL17A is involved in bone loss and postmenopausal osteoporosis^75,76^. Our data shows that the well-established osteo-immune link^77,78^ could play a role in normal variation of skeletal mineralization.

Next, we performed physical variant-to-gene mapping in hMSC-derived osteoblasts, particularly important in the context of bone given that publically available genomic resources typically lack bone data. Using a previously successful approach for identifying target genes at known aBMD-associated loci^15^, we identified three loci for functional follow-up. After siRNA knockdown of 12 genes (4 at each locus), we observed reduced osteoblastic activity and/or reduced mineralization for one gene at each locus (*PRPF38A, KARS* and *TEAD4*, each among the top 70% of expressed genes in hMSC-derived osteoblasts). Two of these genes, *PRPF38A* and *KARS*, are novel in the context of bone. *KARS* encodes the multifunctional protein lysyl-tRNA synthetase, which catalyzes the attachment of amino acids to their cognate tRNAs, but also acts as a signaling molecule when secreted and induces dendritic cell maturation via the MAPK and NFkB pathways^79,80^. On the other hand, *TEAD4* interacts with WWTR1/TAZ transcription co-activators that allow cells to escape negative regulation by the Hippo signaling pathway and undergo increased cell proliferation, the epithelial-mesenchymal transition, and expression of proteins that directly regulate cell adhesion^81^. Osteoblast differentiation is a multi-step process involving the integration of multiple signaling factors, each with its own critical role, and future studies are warranted to dissect which signals are affected by *PRPF38A, KARS*, and *TEAD4* silencing.

Knockdown of *PRPF38A* induced a dramatic morphological change in both hMSC-derived osteoblasts and hFOBs, concurrent with increased expression of adipogenic transcription factors *PPARG* and *FABP4*, suggesting that gene-targeted cells may favor adipogenic differentiation. Little is known about *PRPF38A* or its encoded protein, likely a member of the spliceosome complex that contains a multi-interface protein-protein interaction domain^82^. We recently reported that knockdown of two pediatric aBMD-associated genes, *ING3* and *EPDR1*, resulted in reduced mineralization and also favored adipogenesis^15^. The tightly controlled MSC lineage commitment to adipocytes or osteoblasts is critical for maintaining bone homeostasis^83^, and has been implicated in conditions with abnormal bone remodeling (with increased bone marrow adiposity in osteoporosis^84,85^ and bone loss conditions^86^). In addition to *PRPF38A, ING3*, and *EPDR1*, previous studies suggested that *TEAD4* may promote adipogenesis^87^ in conjunction with *WWTR1*/*TAZ* (signal 5), and that *TGFB* (signal 21) induces a switch from adipogenic to osteogenic differentiation in hMSCs^88^. Further studies are warranted to fully explore the hypothesis that adipogenic vs osteogenic differentiation is a key feature of pediatric bone accrual.

In conclusion, we leveraged a longitudinal modeling approach to both maximize the data available in our cohort and to investigate the genetic determinants of pediatric bone accrual. Our findings suggest that differences in bone accrual attributable to genetic variation are a mechanism linking several of our loci with established associations with later-life fracture risk^6^. Finally, we identified two novel candidate effector genes at two associated loci with no obvious leads and resolved a functional candidate gene among several possible genes at a third locus. At *PRPF38A*, our data strongly supports a putative causal candidate variant, which falls into binding motifs for two relevant transcription factors. Our findings implicate multiple biological pathways involved in variation in bone accrual, and highlight the switch between osteogenesis and adipogenesis as potentially critical in pediatric bone accrual. In conclusion, in-depth longitudinal phenotyping plus appropriate functional follow-up of GWAS loci can yield greater insight into dynamic, complex traits such as bone accrual.

## Methods

### Study cohorts

The BMDCS was a longitudinal, multiethnic cohort of healthy children and adolescents who were recruited from five clinical sites across the United States (Philadelphia, PA; Cincinnati, OH; Omaha, NB; Los Angeles, CA; and New York, NY) to establish aBMD and BMC reference curves^16^. Briefly, the participants were aged 6-16 years at baseline (2002-2003) and were followed for up to 6 additional annual visits (for a maximum total of 7 visits). Older (age 19 yrs) and younger (age 5 yrs) participants were subsequently recruited in 2006-2007 and were followed annually for 2 years (maximum 3 visits). 1885 participants (52% female) had both phenotypes and genetic data, and were thus eligible for inclusion in the present study. Participants 18 years and older gave written informed consent. Parental or guardian consent plus participant assent were obtained for subjects younger than 18 years old. The study was approved by the Institutional Review Board of each respective clinical center.

ALSPAC^89,90^ is a prospective birth cohort study that recruited all pregnant women residing within the catchment area of 3 National Health Service authorities in southwest England with an expected date of delivery between April 1991 and December 1992. In total, 15,454 eligible pregnancies were enrolled in ALSPAC (75% response), with 14,901 livebirths alive at age 1 year. Detailed information has been collected from offspring and parents using questionnaires, data extraction from medical records, linkage to health records, and dedicated clinic assessments up to the last completed contact in 2018. Details of all available data can be found in the ALSPAC study website (http://www.bristol.ac.uk/alspac/researchers/our-data/), which includes a fully searchable data dictionary and variable search tool. Ethics approval was obtained from the ALSPAC law and ethics committee and local research ethics committees. Written informed consent was obtained from all participants. For this study, analysis was performed in white participants (>98% of the sample).

### aBMD and BMC measurement

In the BMDCS, DXA scans of the whole body, lumbar spine, hip, and 1/3 distal radius were obtained using bone densitometers (Hologic, Bedford, MA, USA) following the manufacturer’s guidelines by trained research technicians. The scans were analyzed by the DXA Core Laboratory (University of San Francisco, San Francisco, CA, USA) using Hologic software (v.12.3) for baseline scans and Apex 2.1 for follow-up scans using the “compare” feature. Scans were adjusted for calibration differences among clinical centers and longitudinal drift. aBMD and BMC Z-scores for age, sex and population ancestry were calculated and adjusted for height Z-scores^91^. For growth modeling, unadjusted aBMD or BMC values were used.

In ALSPAC, all participants were invited to undergo up to 6 whole-body DXA scans as part of research clinic assessments at mean ages 9.8, 11.7, 13.8, 15.4, 17.8, and 24.5 years. Scans were performed using a Lunar Prodigy scanner (Lunar Radiation Corp) and were analyzed according to the manufacturer’s standard scanning software and positioning protocols. Scans were reanalyzed as necessary to ensure optimal placement of borders between adjacent subregions, and scans with anomalies were excluded. From these whole-body DXA scans, we extracted BMD and BMC at each age for TBLH and skull.

### Longitudinal modeling of bone accretion

SITAR was used to model individual growth curves separately by sex and ancestry^17^. SITAR is a shape invariant model that generates a mean curve for all included measurements. Individual curves are then described relative to the mean curve by shifting in three dimensions: up-down on the y-axis (differences in mean size, i.e. bone density or content, between subjects relative to the population mean, *a-size*), left-right on the x-axis (differences in age when the rate of growth increases, *b-timing*), and stretched-compressed on the age scale to represent distance over time (how quickly growth occurs, or differences in the rate of bone mineralization in the context of the current study, *c-velocity*). These are estimated as random effects that summarize the difference of each individual growth curve relative to the population mean.

In BMDCS, we performed growth modeling on height and aBMD and BMC measured at the spine, total hip, femoral neck, distal 1/3 radius, skull, and total body less head (TBLH), as previously described^18^. Modeling was performed on up to 2014 children and adolescents (50.7% female and 23.8% self-reported as African American) with a mean of 5 annual study visits each, representing ∼11,000 scans in total (**Supplementary Table 1; Supplementary Figure 1**). Only participants with genetic data and phenotypes were taken forward for heritability and GWAS analyses (max N = 1,399, 51% female, 25% African American).

In ALSPAC, SITAR models were fitted for individuals with at least 1 measurement and were fitted in males and females separately. Initially, the models were fitted to the ALSPAC data alone, and then again with the BMDCS data added. For the combined analyses, fixed effects were included in the model to distinguish between the two cohorts. We were only able to achieve converged models for both sexes for TBLH BMC and skull BMC while modelling both cohorts together. The random-effects (a, b and c) from the fitted models for TBLH and skull BMC (both sexes) were extracted for the ALSPAC participants, converted to sex-specific z-scores, and taken forward for GWAS replication. We included data from 6,382 participants (50% female) (**Supplementary Figure 10**).

### Genotyping and imputation

In BMDCS, genome-wide genotyping was carried out on the Illumina Infinium Omni Express plus Exome BeadChip (Illumina, San Diego, CA) at the Children’s Hospital of Philadelphia Center for Applied Genomics^92^. Quality control was subsequently performed to exclude samples with incorrect or ambiguous gender and with missingness per person >5%, and to exclude variants with call rate <95% and minor allele frequency (MAF) <0.5%. Imputation was performed against the 1000 Genomes Phase 1 v.3 reference panel as previously described^9^. After imputation, variants with MAF < 5% or quality score (INFO) < 0.4 were excluded, yielding 7,238,679 SNPs.

ALSPAC children were genotyped using the Illumina HumanHap550 quad chip genotyping platform (Illumina) by 23andme subcontracting the Wellcome Trust Sanger Institute (Cambridge, UK) and the Laboratory Corporation of America (Burlington, NC, USA). The resulting raw genome-wide data were subjected to standard quality control methods. Individuals were excluded on the basis of gender mismatches; minimal or excessive heterozygosity; disproportionate levels of individual missingness (>3%) and insufficient sample replication (IBD < 0.8). All individuals with non-European ancestry were removed. SNPs with a minor allele frequency of < 1%, a call rate of < 95% or evidence for violations of Hardy-Weinberg equilibrium (P < 5×10^−7^) were removed. Cryptic relatedness was measured as proportion of identity by descent (IBD > 0.1). Related subjects that passed all other quality control thresholds were retained during subsequent phasing and imputation. 9,115 subjects and 500,527 SNPs passed these quality control filters. Of these, we combined 477,482 SNP genotypes in common between the sample of ALSPAC children and ALSPAC mothers. We removed SNPs with genotype missingness above 1% due to poor quality (11,396 SNPs removed) and removed a further 321 subjects due to potential ID mismatches. We estimated haplotypes using ShapeIT (v2.r644) which utilises relatedness during phasing. The phased haplotypes were then imputed to the Haplotype Reference Consortium (HRCr1.1, 2016) panel. The HRC panel was phased using ShapeIt v2, and the imputation was performed using the Michigan imputation server. This gave 8,237 eligible children with available genotype data after exclusion of related subjects using cryptic relatedness measures described previously.

### Heritability analyses

For the SNP heritability analyses, imputed genotypes were converted to ‘best-guess’ genotypes, meaning the genotype call most likely to be true given the imputation dosages, using PLINK with a ‘hard-call’ threshold of 0.499. In PLINK, duplicate SNPs were removed, as well as SNPs with Hardy-Weinberg Equilibrium (HWE) *P* <1×10^−6^ and MAF < 2.5×10^−5^. Additionally, PLINK was used to perform a second round of filtering of SNPs with a missingness rate > 5% and individuals missing genotypes at >5% of SNPs. One of each pair of individuals with an estimated genetic relationship of >0.025 was removed to reduce bias from cryptic relatedness.

Genetic restricted maximum likelihood (GREML)^93^ was used to calculate the amount of trait variance explained by genotyped and imputed SNPs. For the cross-sectional analyses, Z-scores for all phenotypes were adjusted for height-for-age Z-score, with the exception of height and skull, which were not adjusted for height Z-scores. Z-scores were further adjusted for age, sex, cohort (longitudinal set or cross-sectional set), collection site (one of five clinical sites), dietary calcium intake, physical activity^94^, and the first 10 genetic PCs to adjust for population substructure. For the SITAR parameters *a-size, b-timing*, and *c-velocity*, GCTA analysis was performed while adjusting for study site and the first 10 genetic PCs, the only covariates that did not change over time. Sensitivity analyses to examine the effect of ancestry were performed modeling ancestral groups together, as well as with the addition and removal of 10 PCs as covariates.

### Genetic correlation across skeletal sites

We performed GCTA bivariate GREML analysis^93^ for cross-sectional phenotypes to estimate the amount of genetic covariance (the genome-wide effect of causal variants that affect multiple traits) between skeletal sites and aBMD and BMC at each individual skeletal site. PhenoSpd^19^was used to run LD Score Regression genetic correlation analysis^95^ of longitudinal phenotypes. Power calculations for the cross-sectional, longitudinal, and genetic correlation estimates can be found in **Supplementary Table 2**. Most cross-sectional genetic correlation comparisons resulted in high SE estimates, reflecting a large variation surrounding the point estimates and thus low degree of confidence in their accuracy; however, taking only estimates with relatively small SE (<0.10), i.e. those that were most precise, all genetic correlations were high (>0.7) and passed a Bonferroni significance threshold adjusting for the number of comparisons (0.05/78 = 0.00064).

### Genome-wide association analysis

In the BMDCS, GWAS was performed for a total of 36 models: the 6 skeletal sites noted above, for each of the 3 SITAR parameters (*a-size, b-timing, c-velocity*), for both aBMD and BMC. GEMMA^96^ was used to create centered relationship matrices, and mixed effects models (Wald test) were run on sex- and ancestry-standardized SITAR parameters adjusted for collection site (max N with phenotypes, genotypes, and covariates = 1362). The results were subsequently filtered for MAF < 0.05, HWE *P* < 1×10^−6^ and imputation quality (INFO) > 0.4.

### Replication

In ALSPAC, we performed GWAS using a linear mixed model in BOLT-LMM v.2.3^97^. This model estimates heritability parameters and the infinitesimal mixed model association statistics. We also included the first 2 principal components. Genotype data were inputted in PLINK binary format. We used a reference map from BOLT-LMM (build hg19) to interpolate genetic map coordinates from SNP physical (base pair) positions. Reference LD scores supplied by BOLT-LMM and appropriate for analyses of European-ancestry samples were used to calibrate the BOLT-LMM statistic. LD scores were matched to SNPs by base pair coordinate.

We extracted all lead SNPs at our 40 prioritized loci and their proxies (r^2^>0.8 based on a European reference using SNiPA, https://snipa.helmholtz-muenchen.de/snipa3/) from the ALSPAC GWAS results, and looked for broad support at *P* < 0.05.

### Functional candidate gene annotation

We extracted all genes and transcripts in the TADs surrounding each sentinel SNP using the TAD pathways pipeline^33^. The protein-coding genes were then functionally annotated for GO terms, KEGG pathways, and OMIM disease association using the Database for Annotation, Visualization and Integrated Discovery (DAVID) v. 6.8 (https://david.ncifcrf.gov/). To search for pan-tissue eQTLs, we extracted all SNPs in LD (r^2^ > 0.8) with our sentinel SNPs. These SNPs were then queried for significant variant-gene eQTLs in all tissues in GTEx v.7^40^. To search for enriched pathways, all genes and transcripts were subjected to TAD pathway analysis^33^.

### ATAC-seq and high-resolution promoter-focused capture C

The ATAC-seq and capture C methods have been previously published^15^. Briefly, we first identified all proxy SNPs in LD (r^2^ = 0.5) with the sentinel GWAS SNPs using raggr (www.raggr.usc.edu) with the following parameters: populations: CEU+FIN+GBR+IBS+TSI; min MAF = 0.001; min r^2^ = 0.5; max r^2^ = 1.0; max distance = 500kb; max Mendelian errors = 1; HWP cutoff = 0; min genotype % = 75; genotype database = 1000 Genomes Phase 3; genome build hg19. We then assessed which of these proxy SNPs and which of the gene promoters baited in our capture C library resided in an open chromatin region in hMSC-derived osteoblasts, by intersecting their genomic positions with those of the ATAC-seq peaks (using the BEDTools function intersectBed with 1bp overlap). Next, we extracted the chromatin loops linking open proxy SNPs and open gene promoters in the hMSC-derived osteoblast capture C dataset (bait-to-bait interactions were excluded). The results were visualized using the UCSC Genome Browser.

### Functional assays in hMSCs

Primary bone-marrow derived hMSCs isolated from healthy donors (age range: 22-29 yrs) were characterized for cell surface expression (CD166+CD90+CD105+/CD36-CD34-CD10-CD11b-CD45-) and tri-lineage differentiation (osteoblastic, adipogenic, and chondrogenic) potential at the Institute of Regenerative Medicine, Texas A&M University. Expansion and maintenance of the cells were carried out using alpha-MEM supplemented with 16.5% fetal bovine serum (FBS) in standard culture conditions by plating cells at a density of 3000 cells/cm^2^.

Experimental knockdown of candidate genes was achieved using DharmaFECT1 transfection reagent (Dharmacon Inc., Lafayette, CO) using sets of 4 ON-TARGETplus siRNAs in three temporally separated independent hMSC donor lines. Following siRNA transfection, the cells were allowed to recover for 2 days in maintenance media and stimulated with BMP2 for additional 3 days in serum-free osteogenic media as previously described^15^. To assess the influence of knockdown on gene expression (RT-qPCR) and early osteoblast differentiation (histochemical ALP staining), cells were harvested at 3 days post BMP2 stimulation. For extracellular matrix calcium deposition, cells were stained with 0.1% Alizarin Red S after 8-10 days post-BMP2 stimulation. Details of the siRNA and RT-qPCR primers are provided in **Supplementary Tables 19-21**.

For quantification of histochemical ALP stain and Alizarin Red staining, multi-well plates were allowed to air-dry and each well was scanned using high-resolution color bright field objective (1.25X) of the Lionheart FX automated microscope (BioTek). For each scanned well, image analysis was performed using Image J software according to the guidelines provided by the National Institute of Health. For histochemical assays, two independent experiments per siRNA per donor line were performed. P-values for differences between groups were calculated using two-way homoscedastic Students’s *t-*tests.

### Functional assays in hFOBs

hFOB1.19 cells were purchased from ATCC (CRL-11372) and grown in a 1:1 mixture of Ham’s F12 Medium and Dulbecco’s Modified Eagle’s Medium containing no phenol red and supplemented with 10% FBS and 0.3mg/mL G418. Cells were maintained at 33.5^°^C using standard culture conditions and differentiated into mature osteoblasts at 39.5^°^C for all experiments. Knockdown of candidate genes using siRNA and histochemical ALP staining were performed using the same conditions used for the hMSC donor lines. CRISPR-Cas9 mediated deletion of the *PRPF38A* proxy SNP (rs34455069) and sentinel GWAS SNP (rs2762826) were achieved using a pooled lentiviral mCherry construct (Addgene, 99154) containing three sgRNAs on each side of the target. Lentiviral infection was confirmed using mCherry/Texas Red microscopy and efficiency was calculated using the Countess II FL (Thermo). SNP deletions were confirmed using multiplexed sequencing of PCR products generated from hFOB1.19 DNA across the CRISPR region for both SNPs. Quantification of ALP staining in hFOB1.19 cells was similar to that used for hMSC donor lines; however, three technical replicates were used, and the plates were photographed and images converted to grayscale and analyzed using Image J software.

## Data Availability

BMDCS data can be applied for and downloaded via the NIH Data and Specimen Hub (https://dash.nichd.nih.gov/). ALSPAC data is also available on request at http://www.bristol.ac.uk/alspac/researchers/access/.

## References

1. Kalkwarf, H. J. et al. Tracking of Bone Mass and Density during Childhood and Adolescence. The Journal of Clinical Endocrinology & Metabolism 95, 1690–1698 (2010).

2. NIH Consensus Development Panel on Osteoporosis Prevention, Diagnosis, and Therapy. Osteoporosis Prevention, Diagnosis, and Therapy. JAMA: The Journal of the American Medical Association 285, 785–795 (2001).

3. Schettler, A. E. & Gustafson, E. M. Osteoporosis prevention starts in adolescence. J Am Acad Nurse Pract 16, 274–282 (2004).

4. Weaver, C. M. et al. The National Osteoporosis Foundation’s position statement on peak bone mass development and lifestyle factors: a systematic review and implementation recommendations. Osteoporos Int 27, 1281–1386 (2016).

5. Rauch, F. & Schoenau, E. Changes in Bone Density During Childhood and Adolescence: An Approach Based on Bone’s Biological Organization. J Bone Miner Res 16, 597–604 (2001).

6. 23andMe Research Team et al. An atlas of genetic influences on osteoporosis in humans and mice. Nature Genetics (2018) doi:10.1038/s41588-018-0302-x.

7. Estrada, K. et al. Genome-wide meta-analysis identifies 56 bone mineral density loci and reveals 14 loci associated with risk of fracture. Nat Genet 44, 491–501 (2012).

8. Kemp, J. P. et al. Identification of 153 new loci associated with heel bone mineral density and functional involvement of GPC6 in osteoporosis. Nature Genetics 49, 1468–1475 (2017).

9. Chesi, A. et al. A Genomewide Association Study Identifies Two Sex-Specific Loci, at SPTB and IZUMO3, Influencing Pediatric Bone Mineral Density at Multiple Skeletal Sites: GWAS IDENTIFIES TWO SEX-SPECIFIC LOCI INFLUENCING PEDIATRIC BMD. J Bone Miner Res 32, 1274–1281 (2017).

10. Kemp, J. P. et al. Phenotypic Dissection of Bone Mineral Density Reveals Skeletal Site Specificity and Facilitates the Identification of Novel Loci in the Genetic Regulation of Bone Mass Attainment. PLoS Genet 10, e1004423 (2014).

11. Medina-Gomez, C. et al. Life-Course Genome-wide Association Study Meta-analysis of Total Body BMD and Assessment of Age-Specific Effects. The American Journal of Human Genetics 102, 88–102 (2018).

12. Mitchell, J. A. et al. Genetics of Bone Mass in Childhood and Adolescence: Effects of Sex and Maturation Interactions. Journal of Bone and Mineral Research 30, 1676–1683 (2015).

13. Cousminer, D. L. et al. Postmenopausal osteoporotic fracture-associated COLIA1 variant impacts bone accretion in girls. Bone 121, 221–226 (2019).

14. Rowley, M. J. & Corces, V. G. Organizational principles of 3D genome architecture. Nat Rev Genet 19, 789–800 (2018).

15. Chesi, A. et al. Genome-scale Capture C promoter interactions implicate effector genes at GWAS loci for bone mineral density. Nat Commun 10, 1260 (2019).

16. Zemel, B. S. et al. Revised Reference Curves for Bone Mineral Content and Areal Bone Mineral Density According to Age and Sex for Black and Non-Black Children: Results of the Bone Mineral Density in Childhood Study. The Journal of Clinical Endocrinology & Metabolism 96, 3160–3169 (2011).

17. Cole, T. J. et al. Using Super-Imposition by Translation And Rotation (SITAR) to relate pubertal growth to bone health in later life: the Medical Research Council (MRC) National Survey of Health and Development. Int J Epidemiol 45, 1125–1134 (2016).

18. McCormack, S. E. et al. Association Between Linear Growth and Bone Accrual in a Diverse Cohort of Children and Adolescents. JAMA Pediatrics 171, e171769 (2017).

19. Zheng, J. et al. PhenoSpD: an integrated toolkit for phenotypic correlation estimation and multiple testing correction using GWAS summary statistics. GigaScience 7, (2018).

20. Nyholt, D. R. A Simple Correction for Multiple Testing for Single-Nucleotide Polymorphisms in Linkage Disequilibrium with Each Other. Am J Hum Genet 74, 765–769 (2004).

21. AOGC Consortium et al. Whole-genome sequencing identifies EN1 as a determinant of bone density and fracture. Nature 526, 112–117 (2015).

22. Bozec, A. et al. Osteoclast size is controlled by Fra-2 through LIF/LIF-receptor signalling and hypoxia. Nature 454, 221–225 (2008).

23. Hong, J.-H. TAZ, a Transcriptional Modulator of Mesenchymal Stem Cell Differentiation. Science 309, 1074–1078 (2005).

24. Karim, Z. et al. NHERF1 Mutations and Responsiveness of Renal Parathyroid Hormone. New England Journal of Medicine 359, 1128–1135 (2008).

25. Kinoshita, A. et al. Domain-specific mutations in TGFB1 result in Camurati-Engelmann disease. Nat Genet 26, 19–20 (2000).

26. Bialek, P. et al. A twist code determines the onset of osteoblast differentiation. Dev. Cell 6, 423–435 (2004).

27. Marchegiani, S. et al. Recurrent Mutations in the Basic Domain of TWIST2 Cause Ablepharon Macrostomia and Barber-Say Syndromes. The American Journal of Human Genetics 97, 99–110 (2015).

28. Yang, D.-C. et al. Twist Controls Skeletal Development and Dorsoventral Patterning by Regulating Runx2 in Zebrafish. PLoS ONE 6, e27324 (2011).

29. Bradley, E. W., Carpio, L. R., van Wijnen, A. J., McGee-Lawrence, M. E. & Westendorf, J. J. Histone Deacetylases in Bone Development and Skeletal Disorders. Physiological Reviews 95, 1359–1381 (2015).

30. Nakatani, T., Chen, T., Johnson, J., Westendorf, J. J. & Partridge, N. C. The Deletion of Hdac4 in Mouse Osteoblasts Influences Both Catabolic and Anabolic Effects in Bone: CATABOLIC AND ANABOLIC EFFECTS OF HDAC4 DELETION IN OSTEOBLASTS. J Bone Miner Res 33, 1362–1375 (2018).

31. Li, S. et al. A Conditional Knockout Mouse Model Reveals a Critical Role of PKD1 in Osteoblast Differentiation and Bone Development. Sci Rep 7, 40505 (2017).

32. Gadi, J. et al. The Transcription Factor Protein Sox11 Enhances Early Osteoblast Differentiation by Facilitating Proliferation and the Survival of Mesenchymal and Osteoblast Progenitors. J. Biol. Chem. 288, 25400–25413 (2013).

33. Way, G. P., Youngstrom, D. W., Hankenson, K. D., Greene, C. S. & Grant, S. F. Implicating candidate genes at GWAS signals by leveraging topologically associating domains. European Journal of Human Genetics 25, 1286–1289 (2017).

34. Levy-Apter, E. et al. Adaptor Protein GRB2 Promotes Src Tyrosine Kinase Activation and Podosomal Organization by Protein-tyrosine Phosphatase ϵ in Osteoclasts. J. Biol. Chem. 289, 36048–36058 (2014).

35. Patterson, V. L. et al. Mouse hitchhiker mutants have spina bifida, dorso-ventral patterning defects and polydactyly: identification of Tulp3 as a novel negative regulator of the Sonic hedgehog pathway. Human Molecular Genetics 18, 1719–1739 (2009).

36. Cakouros, D. et al. Specific functions of TET1 and TET2 in regulating mesenchymal cell lineage determination. Epigenetics & Chromatin 12, 3 (2019).

37. Ramasamy, S. K., Kusumbe, A. P., Wang, L. & Adams, R. H. Endothelial Notch activity promotes angiogenesis and osteogenesis in bone. Nature 507, 376–380 (2014).

38. Miller, C. H. et al. RBP-J–Regulated miR-182 Promotes TNF-α–Induced Osteoclastogenesis. J.I. 196, 4977–4986 (2016).

39. Almeida, M. Unraveling the role of FoxOs in bone—Insights from mouse models. Bone 49, 319–327 (2011).

40. GTEx Consortium et al. Using an atlas of gene regulation across 44 human tissues to inform complex disease- and trait-associated variation. Nat Genet 50, 956–967 (2018).

41. Levi, G. et al. Defective bone formation in Krox-20 mutant mice. Development 122, 113–120 (1996).

42. Krüger, I. et al. Sp1/Sp3 compound heterozygous mice are not viable: Impaired erythropoiesis and severe placental defects. Dev. Dyn. 236, 2235–2244 (2007).

43. Göllner, H., Dani, C., Phillips, B., Philipsen, S. & Suske, G. Impaired ossification in mice lacking the transcription factor Sp3. Mechanisms of Development 106, 77–83 (2001).

44. Zhao, H. et al. Foxp1/2/4 regulate endochondral ossification as a suppresser complex. Developmental Biology 398, 242–254 (2015).

45. Kotake, S. et al. IL-17 in synovial fluids from patients with rheumatoid arthritis is a potent stimulator of osteoclastogenesis. J. Clin. Invest. 103, 1345–1352 (1999).

46. Geneviève, D. et al. Thromboxane synthase mutations in an increased bone density disorder (Ghosal syndrome). Nature Genetics 40, 284–286 (2008).

47. Shinohara, M. et al. Tyrosine Kinases Btk and Tec Regulate Osteoclast Differentiation by Linking RANK and ITAM Signals. Cell 132, 794–806 (2008).

48. Gao, Y. et al. Estrogen prevents bone loss through transforming growth factor signaling in T cells. Proceedings of the National Academy of Sciences 101, 16618–16623 (2004).

49. Morón, F. J. et al. Multilocus analysis of estrogen-related genes in Spanish postmenopausal women suggests an interactive role of ESR1, ESR2 and NRIP1 genes in the pathogenesis of osteoporosis. Bone 39, 213–221 (2006).

50. Yang, D., Guo, J., Divieti, P., Shioda, T. & Bringhurst, F. R. CBP/p300-Interacting Protein CITED1 Modulates Parathyroid Hormone Regulation of Osteoblastic Differentiation. Endocrinology 149, 1728–1735 (2008).

51. Vega, R. B. et al. Histone Deacetylase 4 Controls Chondrocyte Hypertrophy during Skeletogenesis. Cell 119, 555–566 (2004).

52. Chu, Y. et al. Tet2 Regulates Osteoclast Differentiation by Interacting with Runx1 and Maintaining Genomic 5-Hydroxymethylcytosine (5hmC). Genomics, Proteomics & Bioinformatics 16, 172–186 (2018).

53. Matsumoto, Y. et al. Reciprocal stabilization of ABL and TAZ regulates osteoblastogenesis through transcription factor RUNX2. Journal of Clinical Investigation 126, 4482–4496 (2016).

54. Bollag, W. B. et al. Deletion of protein kinase D1 in osteoprogenitor cells results in decreased osteogenesis in vitro and reduced bone mineral density in vivo. Molecular and Cellular Endocrinology 461, 22–31 (2018).

55. Nesbit, M. A. et al. Mutations Affecting G-Protein Subunit α_11_ in Hypercalcemia and Hypocalcemia. N Engl J Med 368, 2476–2486 (2013).

56. Zhao, X., Chen, L. & Wang, Z. Aesculin modulates bone metabolism by suppressing receptor activator of NF-κB ligand (RANKL)-induced osteoclastogenesis and transduction signals. Biochemical and Biophysical Research Communications 488, 15–21 (2017).

57. Tsurusaki, Y. et al. De novo SOX11 mutations cause Coffin–Siris syndrome. Nat Commun 5, 4011 (2014).

58. Kim, B.-J. et al. Osteoclast-secreted SLIT3 coordinates bone resorption and formation. Journal of Clinical Investigation 128, 1429–1441 (2018).

59. Andrade, A. C., Jee, Y. H. & Nilsson, O. New Genetic Diagnoses of Short Stature Provide Insights into Local Regulation of Childhood Growth. Horm Res Paediatr 88, 22–37 (2017).

60. Grant, S. F. et al. Reduced bone density and osteoporosis associated with a polymorphic Sp1 binding site in the collagen type I alpha 1 gene. Nat. Genet. 14, 203–205 (1996).

61. Uitterlinden, A. G. et al. Relation of Alleles of the Collagen Type Iα1 Gene to Bone Density and the Risk of Osteoporotic Fractures in Postmenopausal Women. New England Journal of Medicine 338, 1016–1021 (1998).

62. Deardorff, M. A. et al. Mutations in Cohesin Complex Members SMC3 and SMC1A Cause a Mild Variant of Cornelia de Lange Syndrome with Predominant Mental Retardation. The American Journal of Human Genetics 80, 485–494 (2007).

63. Nebert, D. W., Wikvall, K. & Miller, W. L. Human cytochromes P450 in health and disease. Phil. Trans. R. Soc. B 368, 20120431 (2013).

64. Somner, J. et al. Polymorphisms in the P450 c17 (17-Hydroxylase/17,20-Lyase) and P450 c19 (Aromatase) Genes: Association with Serum Sex Steroid Concentrations and Bone Mineral Density in Postmenopausal Women. The Journal of Clinical Endocrinology & Metabolism 89, 344–351 (2004).

65. Kushwaha, P., Wolfgang, M. J. & Riddle, R. C. Fatty acid metabolism by the osteoblast. Bone 115, 8–14 (2018).

66. Högström, M., Nordström, P. & Nordström, A. n-3 Fatty acids are positively associated with peak bone mineral density and bone accrual in healthy men: the NO2 Study. The American Journal of Clinical Nutrition 85, 803–807 (2007).

67. Alonso-Pérez, A. et al. Role of Toll-Like Receptor 4 on Osteoblast Metabolism and Function. Front. Physiol. 9, 504 (2018).

68. Kim, H.-N., Iyer, S., Ring, R. & Almeida, M. The Role of FoxOs in Bone Health and Disease. in Current Topics in Developmental Biology vol. 127 149–163 (Elsevier, 2018).

69. Bonjour, J.-P. Calcium and Phosphate: A Duet of Ions Playing for Bone Health. Journal of the American College of Nutrition 30, 438S–448S (2011).

70. Yang, W. et al. The emerging role of Hippo signaling pathway in regulating osteoclast formation. J Cell Physiol 233, 4606–4617 (2018).

71. Van Bezooijen, R. L., Farih-Sips, H. C. M., Papapoulos, S. E. & Löwik, C. W. G. M. Interleukin-17: A New Bone Acting Cytokine In Vitro. Journal of Bone and Mineral Research 14, 1513–1521 (1999).

72. Huang, H. et al. IL-17 stimulates the proliferation and differentiation of human mesenchymal stem cells: implications for bone remodeling. Cell Death Differ 16, 1332–1343 (2009).

73. Croes, M. et al. Proinflammatory T cells and IL-17 stimulate osteoblast differentiation. Bone 84, 262–270 (2016).

74. Wang, Z. et al. IL-17A Inhibits Osteogenic Differentiation of Bone Mesenchymal Stem Cells via Wnt Signaling Pathway. Med Sci Monit 23, 4095–4101 (2017).

75. Won, H. Y. et al. Prominent Bone Loss Mediated by RANKL and IL-17 Produced by CD4+ T Cells in TallyHo/JngJ Mice. PLoS ONE 6, e18168 (2011).

76. Molnár, I., Bohaty, I. & Somogyiné-Vári, É. IL-17A-mediated sRANK ligand elevation involved in postmenopausal osteoporosis. Osteoporos Int 25, 783–786 (2014).

77. Takayanagi, H. et al. T-cell-mediated regulation of osteoclastogenesis by signalling cross-talk between RANKL and IFN-γ. Nature 408, 600–605 (2000).

78. Takayanagi, H. Osteoimmunology and the effects of the immune system on bone. Nature Reviews Rheumatology 5, 667–676 (2009).

79. Lim, H. X. et al. Lysyl–Transfer RNA Synthetase Induces the Maturation of Dendritic Cells through MAPK and NF-κB Pathways, Strongly Contributing to Enhanced Th1 Cell Responses. J.I. 201, 2832–2841 (2018).

80. Park, S. G. et al. Human lysyl-tRNA synthetase is secreted to trigger proinflammatory response. Proceedings of the National Academy of Sciences 102, 6356–6361 (2005).

81. Qi, Y. et al. A splicing isoform of TEAD4 attenuates the Hippo–YAP signalling to inhibit tumour proliferation. Nat Commun 7, ncomms11840 (2016).

82. Schütze, T. et al. Multiple protein–protein interactions converging on the Prp38 protein during activation of the human spliceosome. RNA 22, 265–277 (2016).

83. Chen, Q. et al. Fate decision of mesenchymal stem cells: adipocytes or osteoblasts? Cell Death Differ 23, 1128–1139 (2016).

84. Meunier, P., Aaron, J., Edouard, C. & VlGNON, G. Osteoporosis and the Replacement of Cell Populations of the Marrow by Adipose Tissue: A Quantitative Study of 84 Iliac Bone Biopsies. Clinical Orthopaedics and Related Research 80, 147–154 (1971).

85. Justesen, J. et al. Adipocyte tissue volume in bone marrow is increased with aging and in patients with osteoporosis. Biogerontology 2, 165–171 (2001).

86. Moerman, E. J., Teng, K., Lipschitz, D. A. & Lecka-Czernik, B. Aging activates adipogenic and suppresses osteogenic programs in mesenchymal marrow stroma/stem cells: the role of PPAR-gamma2 transcription factor and TGF-beta/BMP signaling pathways. Aging Cell 3, 379–389 (2004).

87. Zhang, W. et al. The TEA domain family transcription factor TEAD4 represses murine adipogenesis by recruiting the cofactors VGLL4 and CtBP2 into a transcriptional complex. J. Biol. Chem. 293, 17119–17134 (2018).

88. van Zoelen, E. J., Duarte, I., Hendriks, J. M. & van der Woning, S. P. TGFβ-induced switch from adipogenic to osteogenic differentiation of human mesenchymal stem cells: identification of drug targets for prevention of fat cell differentiation. Stem Cell Research & Therapy 7, 123 (2016).

89. Fraser, A. et al. Cohort Profile: The Avon Longitudinal Study of Parents and Children: ALSPAC mothers cohort. International Journal of Epidemiology 42, 97–110 (2013).

90. Boyd, A. et al. Cohort Profile: The ‘Children of the 90s’—the index offspring of the Avon Longitudinal Study of Parents and Children. International Journal of Epidemiology 42, 111–127 (2013).

91. Zemel, B. S. et al. Height Adjustment in Assessing Dual Energy X-Ray Absorptiometry Measurements of Bone Mass and Density in Children. The Journal of Clinical Endocrinology & Metabolism 95, 1265–1273 (2010).

92. Hakonarson, H. et al. A Novel Susceptibility Locus for Type 1 Diabetes on Chr12q13 Identified by a Genome-Wide Association Study. Diabetes 57, 1143–1146 (2008).

93. Yang, J. et al. Common SNPs explain a large proportion of the heritability for human height. Nat Genet 42, 565–569 (2010).

94. Lappe, J. M. et al. The Longitudinal Effects of Physical Activity and Dietary Calcium on Bone Mass Accrual Across Stages of Pubertal Development: PHYSICAL ACTIVITY AND CALCIUM EFFECTS ON BMC ACCRUAL. Journal of Bone and Mineral Research 30, 156–164 (2015).

95. Bulik-Sullivan, B. et al. An atlas of genetic correlations across human diseases and traits. Nature Genetics 47, 1236–1241 (2015).

96. Zhou, X. & Stephens, M. Genome-wide efficient mixed-model analysis for association studies. Nat Genet 44, 821–824 (2012).

97. Loh, P.-R. et al. Efficient Bayesian mixed-model analysis increases association power in large cohorts. Nat Genet 47, 284–290 (2015).

